# The diagnostic value of hyperlipidemia and lipophagy-related genes, PLAUR, IVNS1ABP, and QKI, in acute myocardial infarction

**DOI:** 10.1101/2024.12.05.24318546

**Authors:** Jun-Hua Zou, Hua-Wei Wang, Jia-Zhi Long, Xiao-Na Yang, Li-Hong Yang, Long-Jun Li, Li-Xing Chen, Ling Dong, Jing Chen, Zhao-Hui Meng, Wen Wan

## Abstract

**Background:** Hyperlipidemia (HLP) may intensify myocardial cell damage by disrupting lipophagy, a pivotal lipid metabolism pathway, thereby heightening the risk of acute myocardial infarction (AMI). This study aims to identify HLP- and lipophagy-associated biomarkers for AMI through a combined transcriptomic and mendelian randomization (MR) approach.

**Methods:** The mRNA expression data for AMI, along with HLP-related genes (HRGs) and lipophagy-related genes (LRGs), were sourced from public databases. Biomarkers were identified by conducting differential expression analysis, weighted gene co-expression network analysis (WGCNA), MR analysis, and receiver operating characteristic (ROC) analysis, complemented by two machine learning algorithms and expression validation. These biomarkers facilitated an investigation into the role of platelet activation-related genes (PARGs) in AMI, with enrichment analysis providing insights into their underlying mechanisms. Finally, reverse transcription quantitative polymerase chain reaction (RT-qPCR) was employed to validate biomarker expression in clinical samples.

**Results:** Three biomarkers exhibited a consistently significant upregulation trend in AMI samples, corroborated by RT-qPCR findings. Notably, PLAUR [Odds ratio (OR) = 1.115, 95% confidence interval (CI): 1.006–1.237, *P* = 0.038] and IVNS1ABP (OR = 1.047, 95% CI: 1.000–1.096, *P* = 0.048) emerged as AMI risk factors, while QKI (OR = 0.946, 95% CI: 0.903– 0.991, *P* = 0.020) was identified as a protective factor. Additionally, PLAUR, QKI, and IVNS1ABP demonstrated robust diagnostic efficacy with area under the curve (AUC) values of 0.773, 0.933, and 0.807, respectively; when integrated into a nomogram, the combined AUC reached 0.924. These genes were enriched in pathways linked to cardiovascular diseases, inflammatory responses, and cellular metabolic processes and appeared actively involved in platelet activation, as indicated by their strong associations with PARGs.

**Conclusion:** In summary, the biomarkers PLAUR, QKI, and IVNS1ABP, connected to HLP and lipophagy, showed a causal relationship with AMI and marked diagnostic potential for predicting AMI risk, offering valuable support for clinical diagnostics and AMI research.

## 1. Introduction

Acute myocardial infarction (AMI), commonly known as a heart attack, results from prolonged and severe myocardial ischemia and subsequent necrosis in affected myocardial regions due to acute coronary artery occlusion. AMI occurs approximately every 40 seconds and is a leading cause of mortality, with over one hundred thousand deaths annually and a 30-day post-AMI mortality rate of 13.6%^1^. Although diagnostic methods for AMI have significantly advanced, the electrocardiogram (ECG) and high-sensitivity cardiac troponin I remain the primary diagnostic criteria. These approaches, however, can be inconclusive, particularly in early-stage AMI, cases with mild myocardial injury, or in distinguishing AMI from other conditions with similar symptoms or biomarker elevations^2^. Given the high morbidity and mortality associated with AMI, and the critical need for accurate and timely diagnosis, research into effective biomarkers for the early detection and pathogenesis of AMI has become an international focus for decades, with ongoing studies aiming to refine diagnostic standards and develop innovative technologies.

Hyperlipidemia (HLP), defined as a disorder of lipid metabolism, has emerged as one of the most prevalent medical conditions globally, with incidence rates rising significantly^3^. Similar to risk factors such as smoking, diabetes, and hypertension, HLP is a potent risk factor for AMI. HLP exacerbates atherosclerotic plaque instability, leading to reduced or obstructed blood flow secondary to coronary artery occlusion. Consequently, AMI treatments, including statin therapy, aim to lower blood lipid levels, thereby stabilizing vulnerable plaques and mitigating risk^4^.

Autophagy serves as a cellular mechanism that degrades damaged proteins and organelles *via* lysosomal pathways. Lipophagy, a selective form of autophagy, targets lipid droplets—dynamic organelles that primarily store triglycerides in adipocytes and hepatocytes—and plays a pivotal role in maintaining lipid and metabolic homeostasis across various tissues and cell types^5,6^. Lipophagy activation provides a protective mechanism against diseases associated with abnormal lipid storage. In macrophages, reduced lipophagy leads to intracellular lipid buildup and foam cell formation, thereby accelerating atherosclerosis progression^7–9^. Consequently, lipophagy may stabilize vulnerable plaques, potentially reducing the risk of AMI. Additionally, research has shown that hyperlipidemia, often induced by high-fat diets, is typically associated with decreased autophagy. Impaired lipophagy, in turn, can contribute to hyperlipidemia by disrupting the degradation of triacylglycerol, cholesterol, and other lipids, whereas enhancing autophagy reduces cellular lipid levels^10^. Thus, identifying biomarkers associated with HLP and lipophagy in AMI is essential for advancing the diagnosis and treatment of AMI.

Mendelian randomization (MR) uses genetic variants, commonly single nucleotide polymorphisms (SNPs), as instrumental variables to infer causal relationships within observational data, effectively bypassing confounding influences and mitigating biases^11^. This method mimics the randomization process of randomized controlled trials (RCTs), where treatment allocation is designed to balance clinical characteristics across groups, reducing confounding risk. MR offers a robust research framework, addressing many limitations inherent in traditional observational research and RCTs. Often more cost-effective and efficient, MR capitalizes on genomic data from large-scale genome-wide association studies (GWAS). Particularly valuable for examining potentially harmful health risk factors unsuitable for ethical clinical trials, MR leverages genetic variations fixed at birth to provide insights into the lifelong effects of these risk factors. Recent research identified five genes associated with neutrophil extracellular traps as causal biomarkers for myocardial infarction (MI) using MR combined with bioinformatics methods^12^. Applying MR to identify biomarkers associated with HLP and lipophagy in AMI could be instrumental in uncovering effective diagnostic targets. This study screened for biomarkers linked to HLP and lipophagy in AMI, examining their diagnostic potential and molecular regulatory mechanisms through bioinformatics and MR analysis based on AMI transcriptomic data from public databases, thereby offering a novel reference for clinical AMI diagnosis and treatment.

## 2. Materials and methods

### 2.1 Data source

All data and materials have been made publicly available at the [the gene expression omnibus (GEO)] and can be accessed at [https://www.ncbi.nlm.nih.gov/geo/query/acc.cgi?acc=GSE60993, https://www.ncbi.nlm.nih.gov/geo/query/acc.cgi]. Two transcriptome datasets related to myocardial infarction (MI) were obtained from the gene expression omnibus (GEO) database (https://www.ncbi.nlm.nih.gov/gds). The GSE60993 dataset was based on the GPL6884 platform and included a total of 33 samples^13^. It combined peripheral blood samples from patients with ST-segment elevation MI (STEMI, n = 7) and non-ST-segment elevation MI (NSTEMI, n = 10) to form a disease sample group (n = 17), while seven peripheral blood samples from healthy individuals serve as controls, making up the training set. The GSE166780 dataset, based on the GPL20795 platform, comprised peripheral blood mononuclear cell samples from 8 patients with AMI and eight normal coronary artery peripheral blood samples (control samples), which were used as the validation set^14^. Additionally, based on previously published literature^15^, 1,462 HLP-related genes (HRGs) were obtained from the genecards database (https://www.genecards.org) by referencing the supplementary material files (S1) in Data Sheet 1.zip. Further, using “Lipophagy” as a keyword in the Molecular Signatures Database (MSigDB) (https://www.gsea-msigdb.org/gsea/msigdb), we obtained a gene set named “REACTOME_LIPOPHAGY,” which included 9 lipophagy-related genes (LRGs): PLIN3, PRKAG2, HSPA8, PRKAB1, PRKAG3, PRKAB2, PLIN2, PRKAA2, and PRKAG1.

### 2.2 Identification of differentially expressed genes (DEGs) and key module genes associated with AMI

Differential expression analysis was performed on the training set to identify DEGs between AMI and control samples, using the limma package (v3.58.1)^16^ with criteria of |log2 Fold change (FC)| > 0.5 and *P* < 0.05. Visualization of these DEGs was accomplished through a volcano plot and a heatmap, created with the ggplot2 (v3.4.4)^17^ and ComplexHeatmap (v2.14.0)^18^ packages, respectively. Next, the WGCNA package (v1.71)^19^ facilitated the construction of a co-expression network within the training set, where AMI and control samples served as clinical trait data for weighted gene co-expression network analysis (WGCNA), aiming to identify key modules associated with AMI. Initially, sample clustering was conducted, and outliers were removed to enhance analysis accuracy. To establish a scale-free network, a scale-free fit index (R^2^) threshold above 0.8 was set, with a target mean connectivity close to zero, guiding the determination of the optimal soft threshold power (β). Subsequently, the co-expression matrix was constructed, using the dynamic tree cutting algorithm with a minimum gene count of 100 per module. The MEDissThres parameter was set to 0.3 to merge similar modules identified by the dynamic tree cutting method. Correlations between each module and AMI were then analyzed using the Pearson correlation, with a heatmap illustrating these associations. Modules exhibiting the highest positive and negative correlations with AMI (|cor| > 0.3 and *P* < 0.05) were designated as key modules, and the genes within them were defined as key module genes.

### 2.3 Selection and enrichment analysis of candidate genes related to lipophagy and HLP

Intersection genes were identified by overlapping DEGs with key module genes *via* the VennDiagram package (v1.7.1)^20^. These DEGs were further intersected with LRGs and HRGs individually to isolate differentially expressed LRGs (DE-LRGs) and differentially expressed HRGs (DE-HRGs). Correlations between these gene sets and intersection genes were calculated, selecting highly correlated genes (|cor| > 0.8 and *P* < 0.05) as candidate genes associated with lipophagy and HLP in AMI for subsequent analysis. Gene ontology (GO) and kyoto encyclopedia of genes and genomes (KEGG) enrichment analyses were conducted to identify potential signaling pathways and biological mechanisms relevant to candidate genes, using the ClusterProfiler (v4.7.1.3)^21^ in conjunction with org.Hs.eg.db (v3.16.0)^22^ (*P* < 0.05). Enrichment analysis results were visualized with the ccgraph package (v0.1.0) (https://github.com/gaospecial/ccgraph).

### 2.4 Selection of instrumental variables (IVs)

Expression quantitative trait loci (eQTL) data fo r candidate genes were selected as exposure factors using the Integrative Epidemiology Unit (IEU) Open GWAS database (https://gwas.mrcieu.ac.uk/). The most recent dataset, “ebi-a-GCST90018877,” which contained 24,172,914 SNPs from 461,823 European samples (cases = 20,917; controls = 440,906), was identified by searching for “Myocardial infarction” in the IEU OpenGWAS database and served as the outcome variable. Subsequently, a two-sample MR analysis was conducted to determine genes with potential causal associations with AMI. The MR analysis relied on three core assumptions: (1) a strong and statistically significant association between instrumental variables (IVs) and exposure; (2) IVs independent of confounding variables; and (3) IVs affecting outcomes solely through the exposure pathway.

The IV screening process was initiated using the extract_instruments function from the TwoSampleMR package (v0.56)^23^, which identified SNPs significantly associated with exposure factors (*P* < 5 × 10^-8^). To exclude SNPs in linkage disequilibrium (LD), parameters were set to r^2^ = 0.001 and kb = 5000, with clumping enabled to retain only independent SNPs, thereby reducing potential confounding. The F-statistic (F = beta2/se2) was calculated to confirm the robustness of SNPs, with SNPs having F > 10 considered sufficiently robust. The harmonise_data function from the TwoSampleMR package was subsequently employed to align effect alleles and effect sizes, generating a set of IVs ready for MR analysis.

### 2.5 Identification of genes causally associated with AMI using MR analysis

MR analysis was conducted using the mr function with five algorithms: MR-Egger^24^, Weighted median^25^, Inverse variance weighted (IVW)^26^, Simple mode^23^, and weighted mode^27^. The robustness of the IVW method in detecting causal relationships enabled the identification of significant causal associations (*P* < 0.05) between candidate genes and AMI. An odds ratio (OR) above 1 indicated a risk factor for AMI, whereas an OR below 1 suggested a protective factor. Results of the MR analysis were depicted through scatter plots, forest plots, and funnel plots for a comprehensive visual representation.

To validate the MR findings, a sensitivity analysis was performed. The mr_heterogeneity function facilitated a heterogeneity test *via* Cochran’s Q test^28^, where *P* > 0.05 indicated no significant heterogeneity. Subsequently, the mr_pleiotropy_test function and the MR-PRESSO package (v1.0)^29^ were employed to examine horizontal pleiotropy, with *P* > 0.05 suggesting no substantial pleiotropic effects. The mr_leaveoneout function was then applied for a Leave-One-Out (LOO) sensitivity test^30^. For further assessment of the candidate genes associated with HLP in AMI, receiver operating characteristic (ROC) analysis was performed on both training and validation sets using the pROC package (v1.18.5)^31^. Genes with an area under the curve (AUC) value above 0.7 were considered to have strong discriminatory ability and were selected for additional analysis. These genes were subsequently incorporated into a Least absolute shrinkage and selection operator (LASSO) cox regression analysis using the glmnet package (v4.1.4)^32^, with parameters set to family = binomial and type.measure = class. A 10-fold cross-validation was conducted to compute error rates across various features, identifying features with the lowest error rates at the minimum lambda value, thus designating them as feature genes 1. Additionally, genes meeting the AUC > 0.7 threshold were used to develop a Random Forest (RF) model through the randomForest package (v4.7-1.1)^33^. The RF model assessed the significance of each gene, ranking them by importance, and was subjected to 10-fold cross-validation, with genes achieving peak accuracy on the cross-validation curve defined as feature genes 2. The intersection of these two sets of feature genes ultimately identified the candidate biomarker.

### 2.6 Identifying biomarkers and constructing nomogram

The expression levels of candidate biomarkers were assessed in both AMI and control samples across the training and validation sets. Biomarkers demonstrating consistent trends and significant expression differences across both datasets were designated as validated biomarkers (*P* < 0.05). In the training set, a nomogram was constructed using the rms package (v6.5.0)^34^ to enhance the diagnostic accuracy of these biomarkers for AMI. This nomogram evaluated the model’s diagnostic capacity by assigning each gene a score, with the cumulative score predicting the AMI incidence rate—where higher scores corresponded to higher predicted incidence. Additionally, a calibration curve was plotted to assess the nomogram’s predictive performance, and an ROC curve was generated to evaluate its discriminatory power between AMI and normal samples.

### 2.7 Functional and annotation analyses

Following biomarker identification, a comprehensive analysis was conducted to elucidate their functional roles. To examine their genetic background, chromosomal locations were visualized using the RCircos package (v1.2.2)^35^. The mRNALocater database (https://wolfpsort.hgc.jp/) was then used to predict the subcellular localization of each biomarker, facilitating insights into their roles in disease pathogenesis and potential drug targets in relevant cells. The biomarkers were subsequently uploaded to the GeneMANIA database (http://genemania.org) to identify functionally related genes. Next, each biomarker was treated as a target gene, and correlation coefficients between the biomarker’s expression level and all other genes in the training set were calculated and ranked. Gene set enrichment analysis (GSEA) was then performed for each biomarker using the ClusterProfiler package, with an adjusted *P* < 0.05 significance threshold. The C2: KEGG gene set from MSigDB served as the background for GSEA. The top 10 significantly enriched pathways associated with each AMI-related gene were visualized using ridge plots generated by the enrichplot package (v1.18.3)^36^.

### 2.8 Evaluating the correlation between platelet activation-related genes (PARGs) and biomarkers

Platelets are pivotal in blood coagulation and thrombus formation, with excessive activation and aggregation upon vascular injury being central to AMI pathogenesis^37^. As such, they are critical targets for AMI prevention and treatment. In a previous study^38^, 94 PARGs were identified, and their differential expression was analyzed between AMI and control samples in the training set. Those PARGs showing significant differences were classified as differentially expressed PARGs (DE-PARGs), with their expression patterns visualized in boxplots created using ggplot2 (*P* < 0.05). Spearman correlation analyses were then conducted with the psych package (v2.2.9)^39^ to further examine relationships between biomarkers and DE-PARGs, with results visualized in a heatmap generated by ggplot2 (|cor| > 0.5 and *P* < 0.05).

### 2.9 Construction of regulatory network

To elucidate the potential molecular mechanisms underlying the biomarkers, transcription factors (TFs) potentially regulating these biomarkers were predicted using the ChIP-X enrichment analysis (ChEA) within the NetworkAnalyst database (https://www.networkanalyst.ca/NetworkAnalyst/). The mRNA-TF network was visualized using NetworkAnalyst’s graphical tools. Additionally, target miRNAs interacting with the biomarkers were predicted *via* the miRWalk database (http://mirwalk.umm.uni-heidelberg.de/), selecting those validated by TargetScan (https://www.targetscan.org/vert_80/) and miRDB (http://www.mirdb.org/). Upstream lncRNAs targeting these miRNAs were then retrieved from the starBase database (https://rnasysu.com/encori/), with screening criteria set to clipExpNum > 6 and pancancerNum > 2. Using this information, a comprehensive lncRNA-miRNA-mRNA regulatory network was constructed and visualized with Cytoscape software.

### 2.10 Disease association analysis and drug prediction studies

To explore associations between biomarkers and other diseases, the DisGeNET database (https://www.disgenet.org/) was used *via* the DisGeNET plugin within Cytoscape software to retrieve disease correlations. Concurrently, drugs targeting these biomarkers were predicted using the Drug SIGnatures database (DSigDB) (https://dsigdb.tanlab.org/DSigDBv1.0/) with an adjusted *P* < 0.05 threshold, and a network diagram was constructed for visualization.

### 2.11 Expression validation of biomarkers

To validate the identified biomarkers, rtranscription quantitative polymerase chain reaction (RT-qPCR) was performed on peripheral blood samples from 5 patients with AMI and 5 healthy controls. All samples were obtained from the First Affiliated Hospital of Kunming Medical University with informed consent from donors. Ethical approval was granted by the Ethics Committee of the First Affiliated Hospital of Kunming Medical University (approval number: 2044L84). Total RNA extraction was performed using the Trizol method (Ambion, 15596-018CN, USA), and cDNA synthesis was conducted with the SweScript First Strand cDNA synthesis kit (Servicebio, G3333-50, China). GAPDH served as an internal reference, and the 2^-ΔΔCt^ method^40^ was applied to calculate biomarker expression levels, with statistical significance determined at *P* < 0.05. Primer sequences are listed in Table S1.

### 2.12 Statistical analysis

All analyses were conducted in R software (v4.2.2), and group differences were assessed *via* the Wilcoxon test, with *P* < 0.05 denoting statistical significance.

## 3. Results

### 3.1 Identification of 1,141 DEGs and 5,572 key module genes associated with AMI

In the training set of AMI and control samples, 1,141 DEGs were identified, with 653 genes upregulated and 488 genes downregulated in AMI samples (Fig. 1A, B). Using AMI and control samples as clinical traits, a co-expression network was constructed. Clustering analysis confirmed adequate clustering across samples, with no outliers detected (Fig. 1C). To achieve a scale-free network structure, a scale-free fit index (R^2^) threshold above 0.8 was set, with mean connectivity close to zero, establishing an optimal soft threshold power (β) of 7 (Fig. 1D). According to the dynamic tree cutting method, a minimum module gene count of 100 was used, resulting in 39 identified modules (Fig. 1E). By setting MEDissThres to 0.3, similar modules were merged, yielding 11 consolidated modules (Fig. 1F, G). A subsequent heatmap illustrated module correlations with clinical traits (AMI and control) (Fig. 1H). Based on selection criteria, the module most positively correlated with AMI (MEmidnightblue; cor = 0.49, *P* < 0.05) and the module most negatively correlated with AMI (MEblue; cor = −0.6, *P* < 0.05) were chosen as key modules. MEmidnightblue contained 1,736 genes, and MEblue contained 3,836 genes, culminating in a total of 5,572 AMI-associated key module genes after merging.

**Figure 1.**
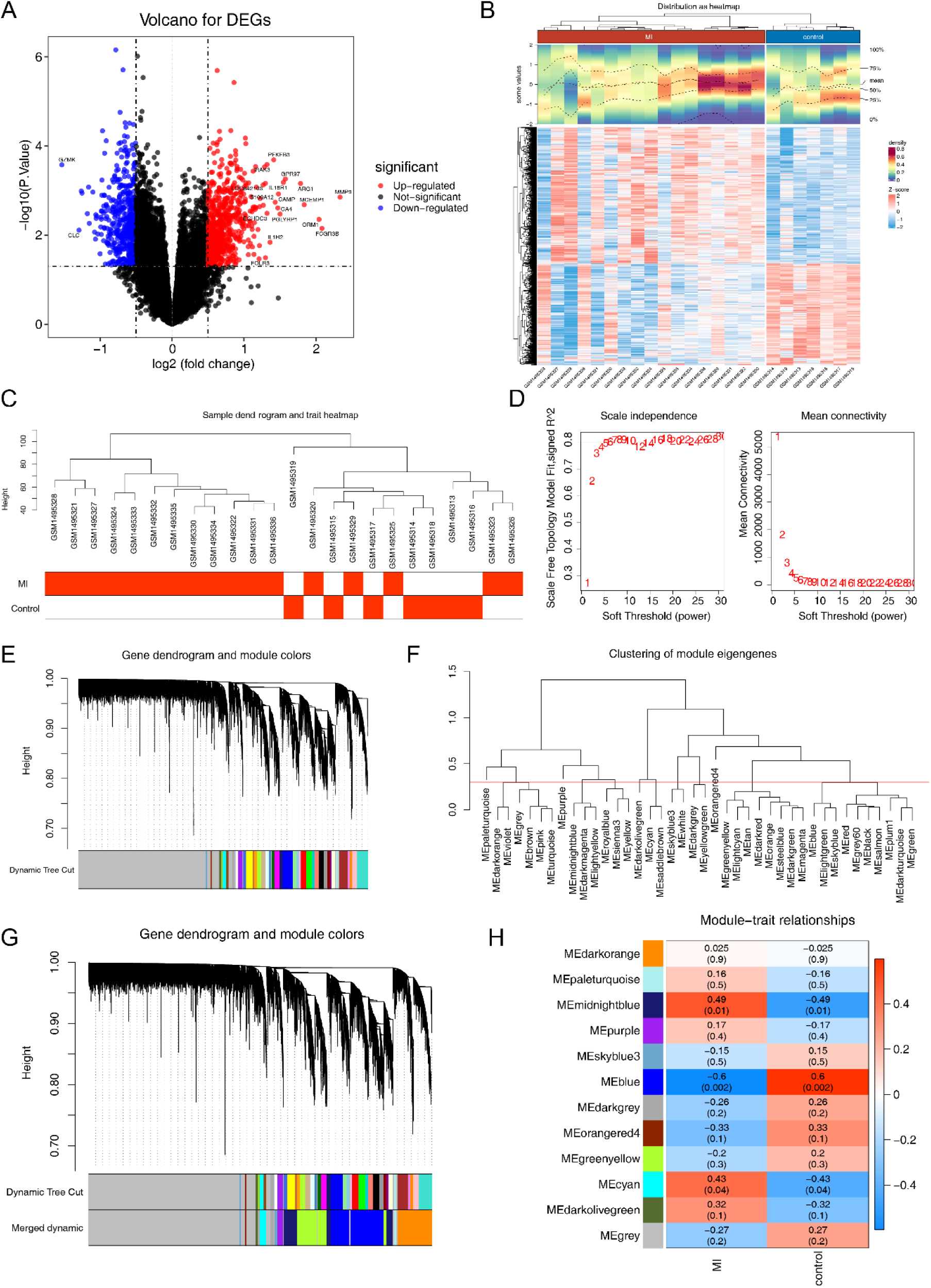
Differential expression analysis and weighted gene co-expression network analysis. (A) Volcano map of differentially expressed genes. (B) Expression heat map of differentially expressed genes. (C) Sample clustering and trait heat map. (D) Scale-free soft threshold distribution. (E) Module clustering dendrogram. (F) Clustering of module eigengenes. (G) Different module genes are merged with traits, and genes are classified into various modules (different colors) by hierarchical clustering, where gray defaults to genes that cannot be classified into any module. (H) Heatmap of module correlation with clinical traits.

### 3.2 Exploring the significance of 917 candidate genes biological functions and cell signaling pathways

By intersecting the 1,141 DEGs with the 5,572 key module genes, alongside 1,462 HRGs and 9 LRGs, 1,011 intersection genes, 110 DE-HRGs, and 1 DE-LRG were identified (Fig. 2A-C).

**Figure 2.**
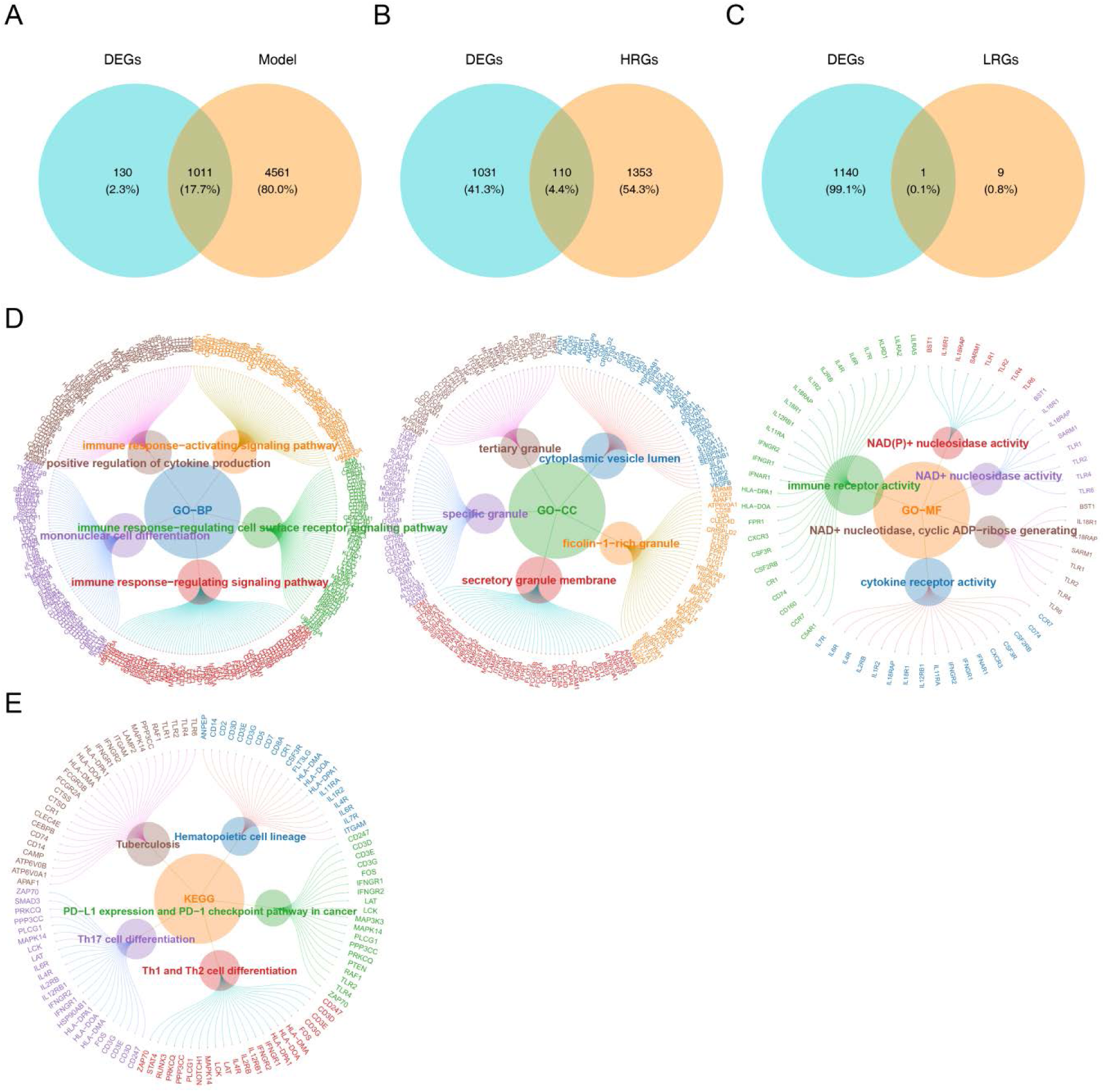
Screening and functional enrichment analysis of candidate genes. (A-C) Veen plots of differential module genes and differential genes. (D) Gene ontology enrichment results of candidate genes. (E) Kyoto encyclopedia of genes and genomes enrichment results of candidate genes.

Upon calculating correlations between the 110 DE-HRGs and 1 DE-LRG with the 1,011 intersection genes, 917 strongly correlated genes were selected as candidate genes (|cor| > 0.8, *P* < 0.05). Enrichment analysis of these 917 candidate genes revealed significant involvement in various biological functions and pathways, identifying 1,011 Gene Ontology-biological process (GO-BP) terms, 72 GO-cellular component (GO-CC) terms, and 102 GO-molecular function (GO-MF) terms. These were primarily linked to immune response and cell differentiation in biological processes, binding activity and receptor/transmembrane signaling in cellular components, and enzyme activity in molecular functions, with additional relevance to secretion and vesicular transport (Fig. 2D). Additionally, 34 KEGG pathways were identified (Fig. 2E), including “tuberculosis,” “hematopoietic cell lineage,” “Th17 cell differentiation,” “Th1 and Th2 cell differentiation,” and “PD-L1 expression and PD-1 checkpoint pathway in cancer,” highlighting associations with immune regulation, cell signaling, enzyme activity, and cellular secretion and transport.

### 3.3 Selection of 31 genes with causal associations to AMI

MR analysis was performed on 917 candidate genes to identify those with causal associations with AMI, using the genes as exposure factors and AMI as the outcome variable. Following IV filtering, 634 eQTL data points were obtained, comprising a total of 2,616 SNPs. Robustness validation using F-statistics refined the IVs to 174 SNPs. MR analysis *via* the IVW algorithm identified 31 genes with causal links to AMI, notably highlighting PLAUR, QKI, and IVNS1ABP. Specifically, PLAUR [OR = 1.115, 95% Confidence Interval (CI): 1.006–1.237, *P* = 0.038] and IVNS1ABP (OR = 1.047, 95% CI: 1.000–1.096, *P* = 0.048) were confirmed as risk factors, while QKI (OR = 0.946, 95% CI: 0.903–0.991, *P* = 0.020) was found to be protective against AMI (Fig. 3). Scatter plots demonstrated significant associations between the 31 genes and AMI (Fig. S1), with forest plots illustrating substantial effect sizes in the IVW model (Fig. S2). Funnel plots further validated adherence to MR principles, supporting consistency with Mendel’s second law of independent assortment (Fig. S3).

**Figure 3.**
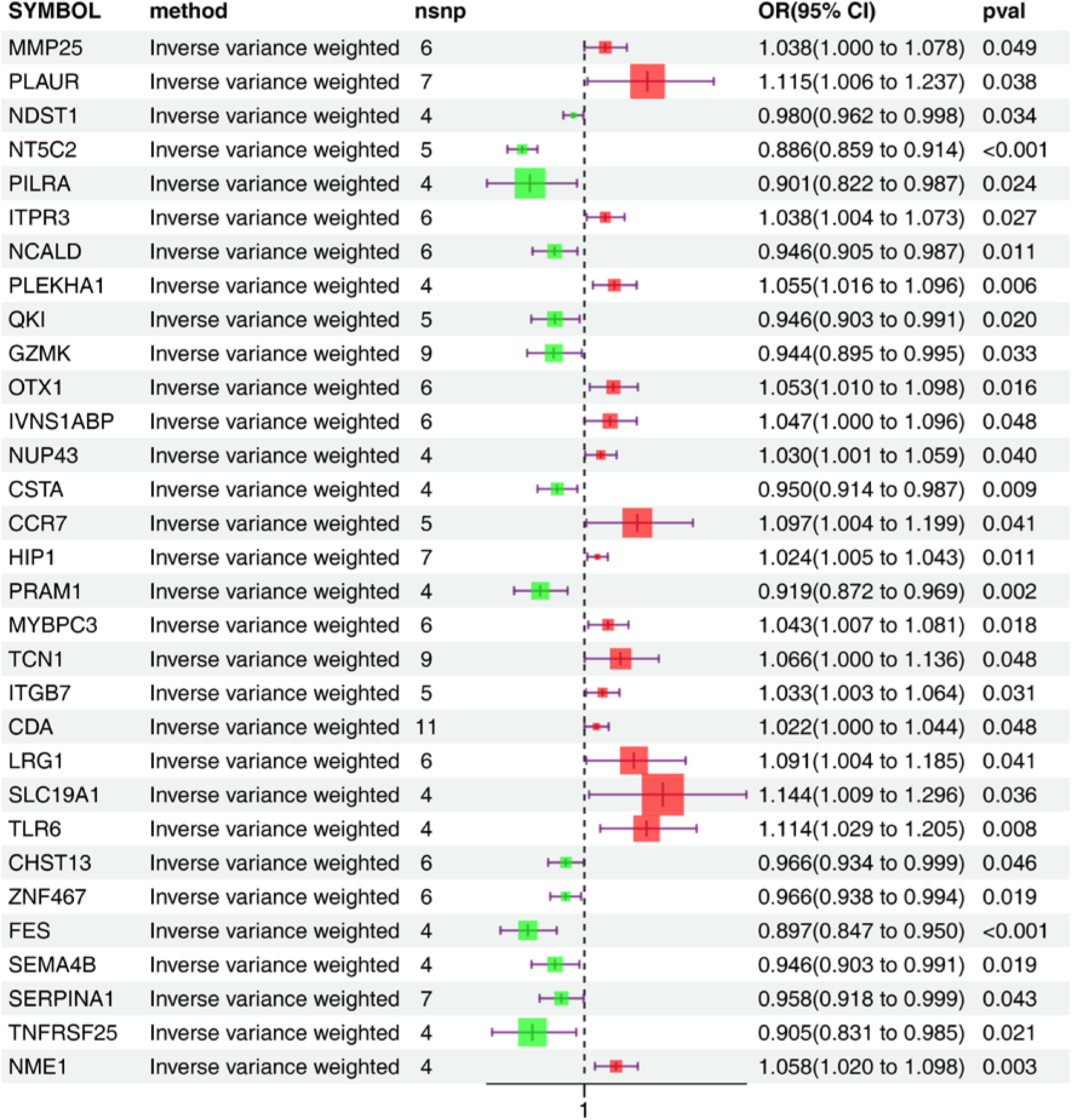
Forest map by inverse variance weighted method.

To reinforce MR findings, a comprehensive sensitivity analysis was conducted. Cochran’s Q test confirmed the lack of heterogeneity influence on PLAUR and FES results under the IVW multiplicative random effects model (*P* < 0.05), allowing all 31 genes to proceed to further analysis (Table S2). Horizontal pleiotropy testing revealed no confounding bias (both mr_heterogeneity and MRPRESSO *P* > 0.05) (Table S3). LOO analysis indicated stability, confirming that no single SNP drove the results (Fig. S4). Finally, the Steiger test verified unidirectional causality from these genes to AMI (correct_causal_direction = true and *P* < 0.05) (Table S4), supporting the directional causal relationship.

### 3.4 Recognition of PLAUR, QKI, OTX1, IVNS1ABP, CHST13, and FES as candidate biomarkers

ROC analysis was conducted to evaluate the diagnostic performance of the 31 genes, revealing AUC values of 0.773, 0.933, and 0.807 for PLAUR, QKI, and IVNS1ABP, respectively, in the training set (Fig. 4A, B). Sixteen genes effectively differentiated AMI from control samples in both training and validation sets and were thus included in subsequent machine learning analyses. LASSO regression identified 9 feature genes (PLAUR, PILRA, QKI, OTX1, IVNS1ABP, CCR7, CHST13, ZNF467, and FES) at the optimal lambda.min of 0.0226, which minimized error rates (Fig. 4C, D). Meanwhile, the RF model demonstrated peak accuracy with 8 selected genes (OTX1, QKI, CHST13, TLR6, FES, ITGB7, IVNS1ABP, and PLAUR), designated as feature genes 2 (Fig. 4E, F). The intersection of these two gene sets yielded 6 candidate biomarkers: PLAUR, QKI, OTX1, IVNS1ABP, CHST13, and FES (Fig. 4G).

**Figure 4.**
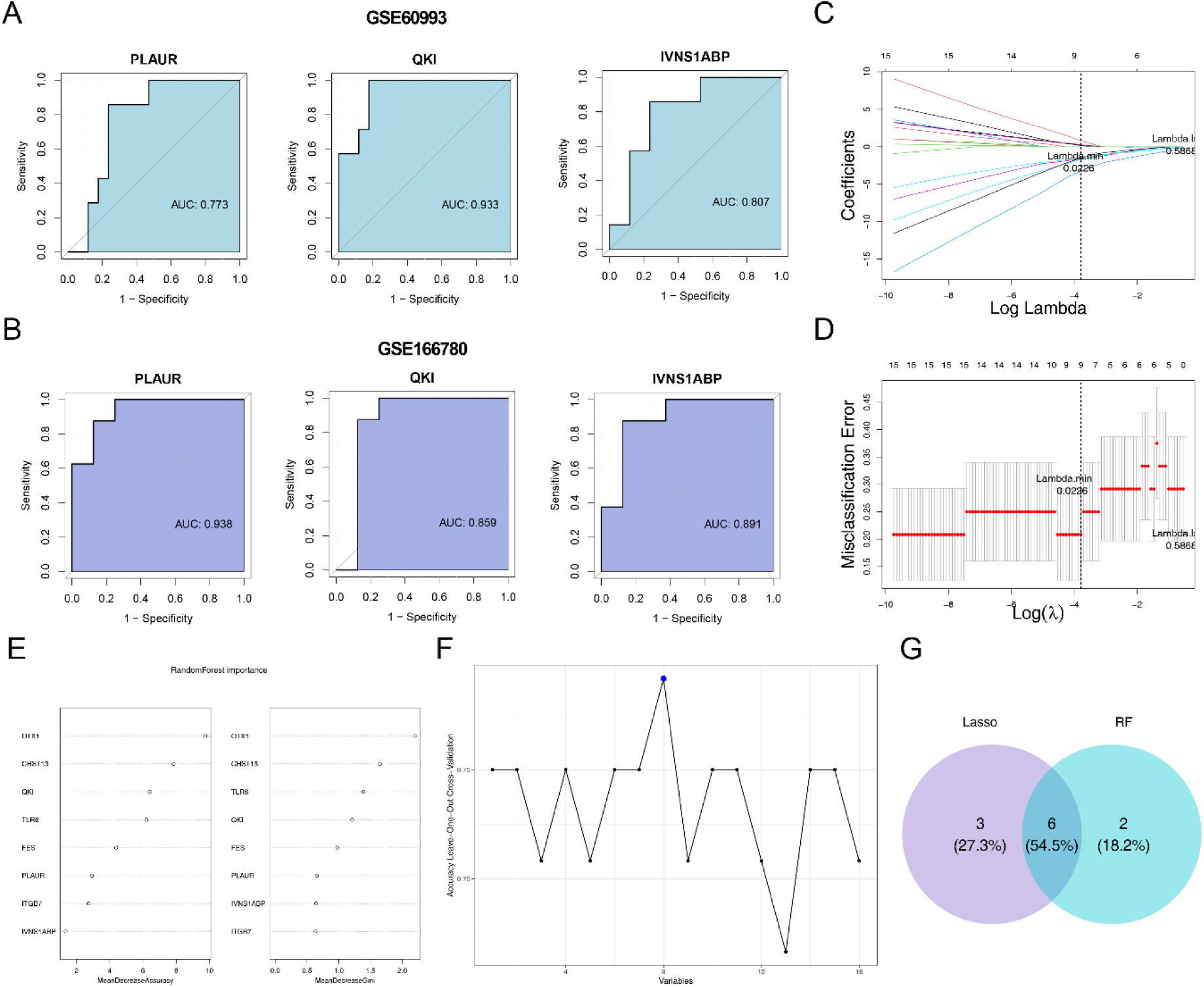
Screening of biomarkers and their diagnostic value. (A) ROC curves for candidate key genes (training set GSE60993). (B) ROC curves for candidate key genes (validation set GSE166780). (C) Plot of gene coefficients versus penalty coefficients. (D) Error plots for cross-validation. (E) Random forest regression model. (F) Cross-validation curves. (G) Machine learning screening of biomarkers for Venn Diagrams. ROC: Receiver operating characteristic; AUC: Area under the curve.

### 3.5 Nomogram constructed using biomarkers PLAUR, QKI and IVNS1ABP demonstrated robust potential

The expression of these 6 candidate biomarkers was then assessed in AMI and control samples across both training and validation sets (Fig. 5A, B). Among them, only PLAUR, QKI, and IVNS1ABP showed consistent upregulation with significant differences (*P* < 0.05) in AMI samples, leading to their selection as key biomarkers associated with lipophagy and HLP in AMI.

**Figure 5.**
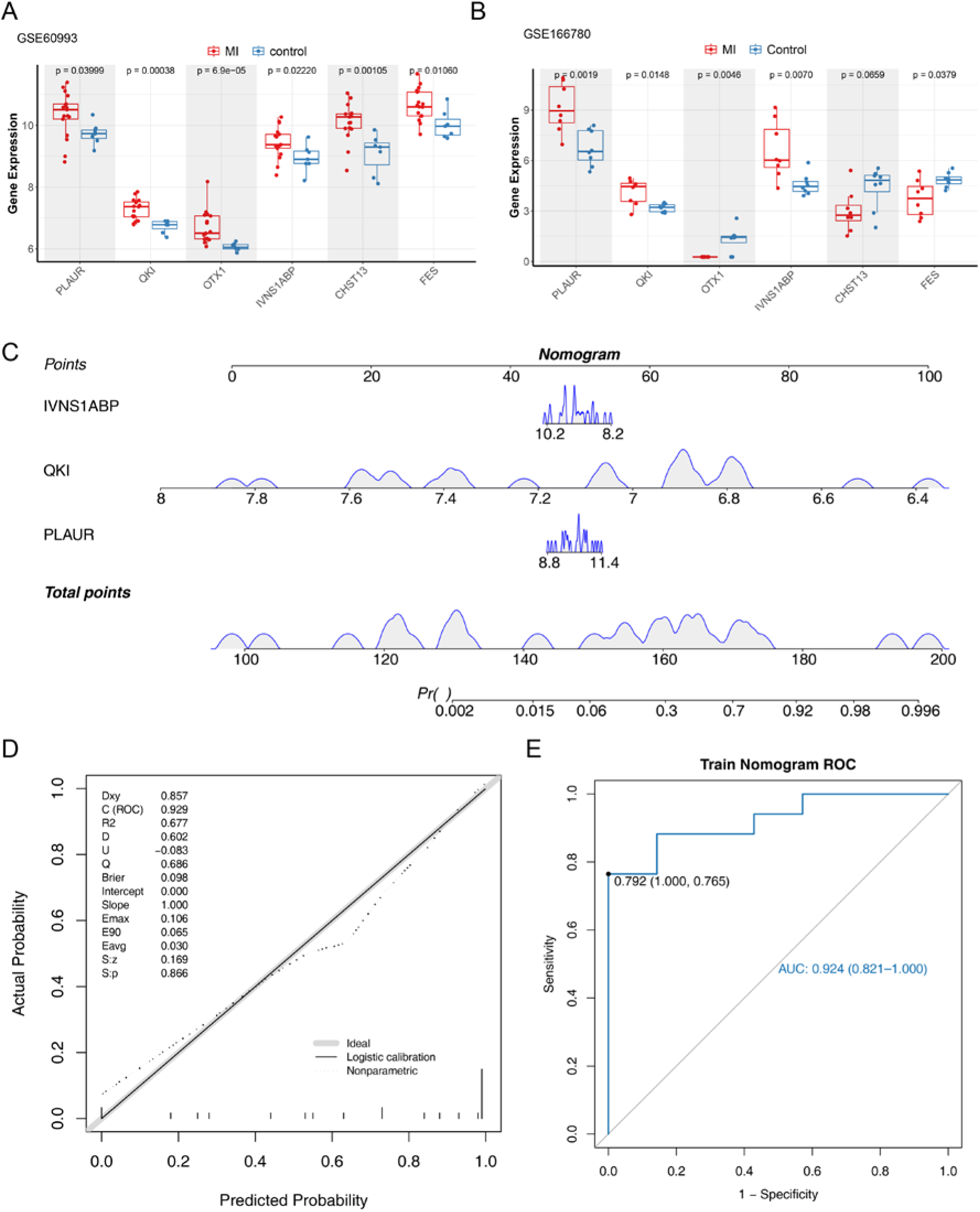
Expression validation of biomarkers and construction of nomogram. (A) Boxplots of the expression of the characterized genes in the GSE60993. (B) Boxplots of the expression of the characterized genes in the GSE166780. (C) Prediction of morbidity by column line graphs. (D) Calibration curves for nomogram. (E) ROC curves for nomogram. ROC: Receiver operating characteristic; AUC: Area under the curve.

Given the strong diagnostic performance and consistent expression trends of these 3 genes, they were incorporated into a nomogram to harness their combined diagnostic potential for clinical applications (Fig. 5C). The calibration curve indicated excellent model performance, closely aligning with the ideal slope of 1 (Fig. 5D). The ROC curve analysis showed an optimal nomogram threshold of 0.792, with specificity and sensitivity of 1.000 and 0.765, respectively. The overall AUC reached 0.924, significantly surpassing the diagnostic potential of PLAUR or IVNS1ABP when used individually (Fig. 5E).

### 3.6 Deciphering functional implications of biomarkers in critical pathways

Chromosomal localization analysis identified IVNS1ABP on chromosome 1, QKI on chromosome 6, and PLAUR on chromosome 19 (Fig. 6A). Subcellular localization revealed predominant nuclear expression for IVNS1ABP and QKI, whereas PLAUR showed maximal cytoplasmic expression (Fig. 6B). The GeneMANIA network further identified 20 genes closely linked to these biomarkers, with PGAP1 displaying strong interactions with all three (Fig. 6C). GSEA indicated that the biomarkers were primarily enriched in pathways such as “lipid and atherosclerosis,” “complement and coagulation cascades,” “FoxO signaling pathway,” “apoptosis,” “platelet activation,” “oxidative phosphorylation,” “HIF-1 signaling pathway,” “TNF signaling pathway,” and “VEGF signaling pathway” (Fig. 6D-F). These pathways are chiefly associated with cardiovascular diseases, inflammatory responses, and cellular metabolic processes, suggesting critical routes through which these biomarkers may impact the onset and progression of AMI.

**Figure 6.**
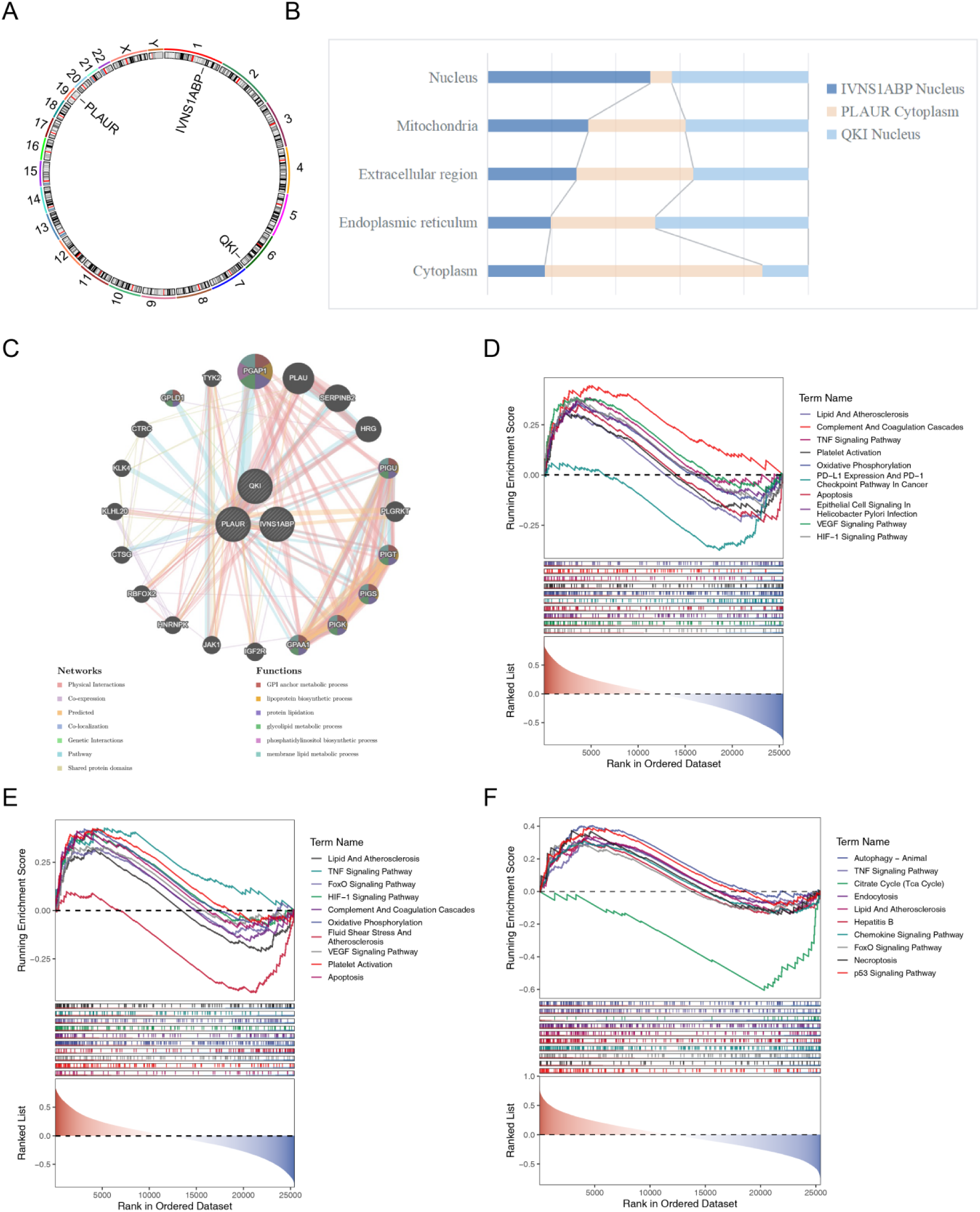
Results of functional analysis of biomarkers. (A) Biomarker chromosome localization circle map. (B) Histogram of subcellular localization of biomarkers. (C) GeneMANIA network. (D) Ridges of GSEA enrichment for the PLAUR. (E) Ridges of GSEA enrichment for the QKI. (F) Ridges of GSEA enrichment for the IVNS1ABP. GSEA: Gene set enrichment analysis.

### 3.7 Deciphering the roles of PARGs in AMI

Further analysis of 94 PARGs in the training set revealed 12 DE-PARGs between AMI and control samples (*P* < 0.05) (Fig. 7A). Notably, genes such as SLC7A11, VAV1, DGKI, P2RX1, and CLIC1 exhibited significantly higher expression in AMI samples (*P* < 0.05), suggesting a potential role in AMI pathogenesis related to platelet activation or aggregation. In contrast, genes including CD40, ENTPD2, CD40LG, FIBP, TSPAN32, DGKE, and LCK showed decreased expression in AMI samples (*P* < 0.05), implying a potential suppression of platelet functionality, which may reduce platelet activity and contribute to vascular inflammation and thrombus formation.

**Figure 7.**
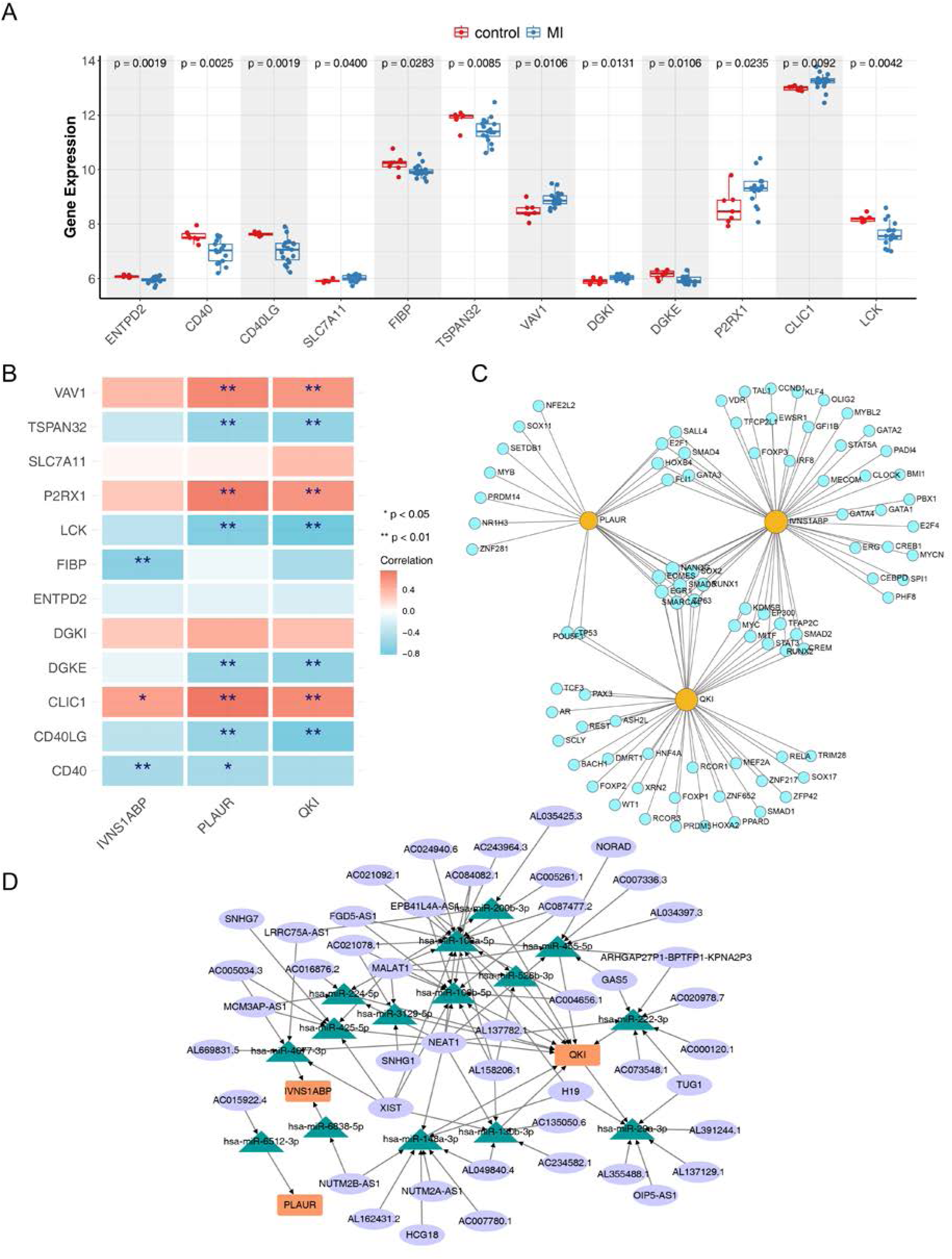
(A) Boxplot of differences in PARGs in normal and diseased samples. (B) Heatmap of biomarker correlation with DE-PARGs. * represents P < 0.05, ** represents P < 0.01, *** represents P < 0.001, **** represents P < 0.0001 (C) mRNA-TF network. The yellow color in the figure represents biomarkers and the blue color represents transcription factors. (D) miRNA-biomarker regulatory network. Orange squares represent biomarkers; green triangles represent miRNAs; purple node circles represent lncRNAs. TF: Transcription factors; PARGs: platelet activation-related genes; DE-PARGs: Differentially expressed PARGs.

Correlation analysis further highlighted strong positive correlations between PLAUR and CLIC1 (cor = 0.76, *P* < 0.05) and between PLAUR and P2RX1 (cor = 0.71, *P* < 0.05), suggesting possible joint involvement in AMI-related processes such as platelet activation or inflammatory responses. In contrast, a strong negative correlation was observed between PLAUR and LCK, as well as between QKI and both CD40LG (cor = −0.77, *P* < 0.05) and LCK (cor = −0.72, *P* < 0.05) (Fig. 7B), indicating that these genes may exert opposing effects on platelet regulation or may be associated with a suppressed platelet state.

### 3.8 Constructing regulatory networks to explore potential mechanisms of biomarkers

Using the NetworkAnalyst database, 84 TFs potentially regulating the three biomarkers were identified, leading to the construction of an mRNA-TF network comprising 87 nodes and 114 edges. Notably, TFs such as NANOG, SOX2, EOMES, SMAD3, RUNX1, EGR1, SMAGCA4, and TP63 were found to interact with all biomarkers simultaneously (Fig. 7C).

Through the miRWalk database, 87 target miRNAs interacting with the three biomarkers were predicted. Following further selection *via* the starBase database, 46 upstream lncRNAs of these miRNAs were identified. These data facilitated the construction of a lncRNA-miRNA-mRNA network, which included 15 miRNAs, 46 lncRNAs, and 3 biomarkers, resulting in 100 complex interactions (Fig. 7D). For instance, lncRNAs such as MALAT1, XIST, NEAT1, and AC024940.6 may regulate QKI *via* hsa-miR-106a-5p.

### 3.9 Exploring the relationship between biomarkers, diseases, and targeted drugs

Disease associations for the biomarkers were retrieved from the DisGeNET database, revealing that IVNS1ABP was related to two diseases, including Prostatic Neoplasms, while QKI was associated with nine diseases, such as Glioma, and PLAUR was linked to twenty diseases, including Inflammation (Fig. 8A). Drug prediction analysis identified 12 potential targeting drugs, with eight targeting IVNS1ABP, six targeting PLAUR, and seven targeting QKI. Notably, piperlongumine and compound 7646-79-9 were found to target all three biomarkers simultaneously (Fig. 8B).

**Figure 8.**
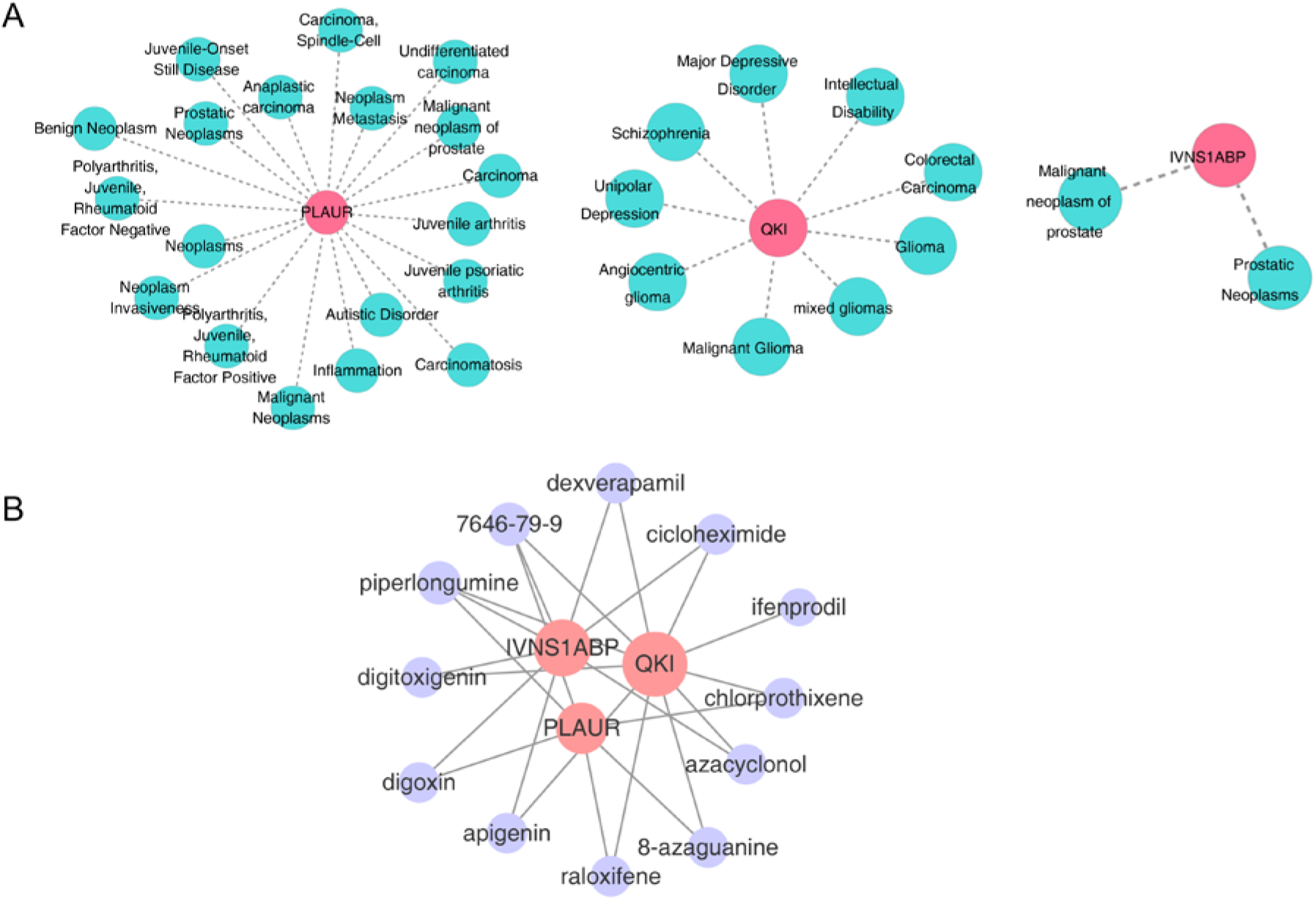
Disease network analysis of biomarkers. (A) Gene-disease networks predicted by the DisGeNET database. Pink nodes represent biomarkers and cyan blue represents diseases. (B) Gene-drug network predicted by DSigDB database. Pink nodes represent biomarkers, purple represents drugs. DSigDB: The drug SIGnatures database.

### 3.10 Verification of biomarkers expression

Previous findings indicated that PLAUR, QKI, and IVNS1ABP were significantly overexpressed in AMI samples (*P* < 0.05) (Fig. 5A, B), prompting further validation through RT-qPCR. RT-qPCR analysis confirmed significantly elevated expression levels for all three biomarkers in AMI samples (*P* < 0.05), consistent with observations from the public dataset (Fig. 9A-C).

**Figure 9.**
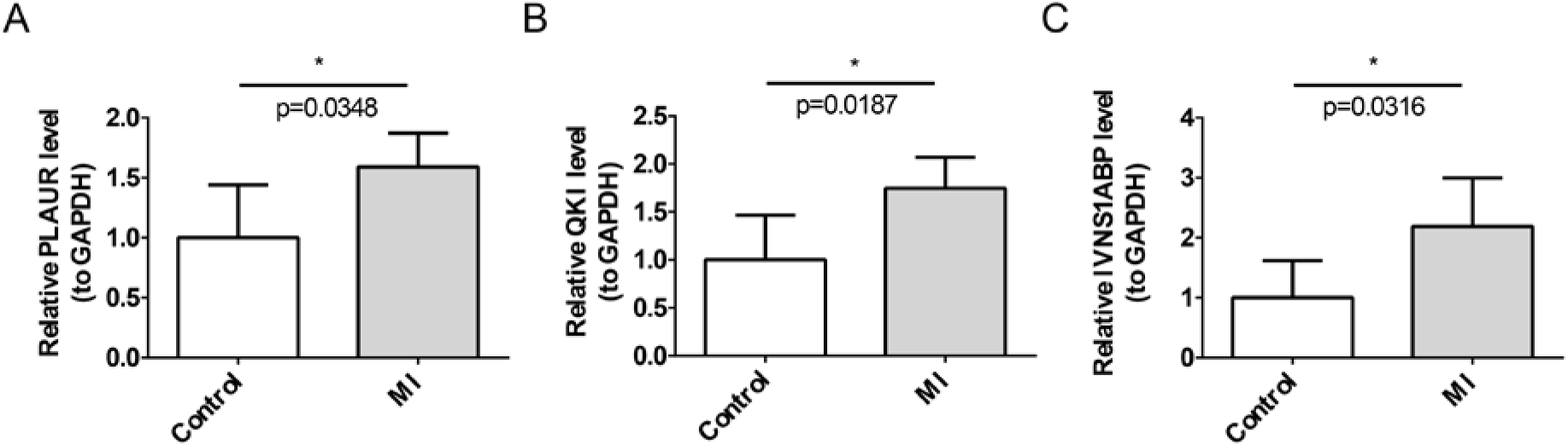
Validation of biomarker expression by RT-qPCR. (A) PLAUR. (B) QKI. (C) IVNS1ABP. * represents P < 0.05. RT-qPCR: Reverse transcription quantitative polymerase chain reaction.

## 4. Discussion

AMI presents a significant public health challenge worldwide, affecting both developed and developing nations with its high morbidity and mortality. Thus, the prompt identification, prevention, diagnosis, and treatment of AMI are essential for reducing global mortality associated with this condition. HLP has been widely recognized as a critical risk factor for AMI, contributing to the destabilization of atherosclerotic plaques^41^. Lipophagy, a selective degradation process targeting lipid droplets, is crucial for maintaining lipid and metabolic homeostasis^7^. However, predictive and diagnostic biomarkers, as well as therapeutic targets related to HLP and lipophagy in the context of AMI, remain largely unexplored. In this study, transcriptomic data from public databases (GSE60993 and GSE166780) were analyzed, identifying AMI-associated biomarkers based on LRGs and HRGs. Furthermore, bioinformatics analyses were conducted to elucidate the molecular regulatory mechanisms of these biomarkers, their roles in platelet activation, disease associations, and potential drug targets.

A total of 917 strongly correlated candidate genes were identified. Although HRGs maintained HLP-related functions and LRGs retained lipophagy-associated roles, this study uncovered additional underlying mechanisms and potential interactions between HLP and lipophagy pathways. Enrichment analyses demonstrated that HLP- and lipophagy-related DEGs were significantly involved in pathways linked to immune responses, enzymatic activities, signaling pathways, regulatory processes, and cellular secretion and transport mechanisms. GO and KEGG enrichment analyses highlighted pathway associations, including “tuberculosis,” “hematopoietic cell lineage,” “PD-L1 expression and the PD-1 checkpoint pathway in cancer,” “Th1 and Th2 cell differentiation,” and “Th17 cell differentiation,” which have not been widely reported in the context of AMI. Hematopoietic cell lineage, especially monocytes and macrophages, plays a pivotal role in lipid metabolism through cytokine and chemokine production. Additionally, macrophages’ uptake of modified lipoproteins can lead to foam cell formation, a hallmark of atherosclerotic plaques. Targeting specific hematopoietic cell populations or their signaling pathways may offer novel strategies for managing HLP and its related cardiovascular risks. Lipophagy may facilitate lipid turnover, balancing glycolysis and oxidative phosphorylation by modulating fatty acid availability, which is vital for the energy demands and membrane integrity of these cells. Furthermore, pathways associated with hematopoietic cell lineages, such as MAPK, PI3K/Akt, and JAK/STAT, are integral to AMI pathophysiology and represent promising therapeutic targets^42^.

The 917 candidate genes were subjected to a two-sample MR analysis, identifying 31 genes with causal relationships to AMI. Notably, PLAUR, QKI, and IVNS1ABP were identified as significant DEGs in both the training and validation sets, with consistent expression trends, suggesting their potential as biomarkers related to lipophagy and HLP in AMI. Combined with MR analysis findings, PLAUR and IVNS1ABP emerged as AMI risk factors, whereas QKI was identified as a protective factor with strong potential.

Quaking (QKI), an RNA-binding protein and member of the STAR family, is abundantly expressed in the heart and other organs, where it plays a critical role in post-transcriptional regulation of mRNA, including splicing, stability, and translation ^43,44^. The anti-apoptotic function of QKI has been linked to its interaction with FoxO1, a transcription factor involved in cell death and growth inhibition. Evidence suggests that QKI suppresses apoptosis in cardiomyocytes, potentially by modulating FoxO1, and that its downregulation in diabetic hearts contributes to increased ischemic vulnerability. Dysregulation of QKI is associated with cardiovascular diseases and has been shown to influence macrophage polarization in endotoxic shock^45–47^. Additionally, QKI interacts with miR-155, whose downregulation by miR-155 has been implicated in promoting MI-induced apoptosis, underscoring QKI’s role in cardiac protection^48^. Consistently, our MR analysis supported QKI as a protective factor against AMI. The Influenza virus NS1A-binding protein (IVNS1ABP), part of the Kelch family, plays a role in actin cytoskeleton organization, essential for cellular functions such as locomotion, division, and phagocytosis. Known for its involvement in responses to viral infections, IVNS1ABP is also implicated in cellular defense mechanisms^49^. Dysregulation of IVNS1ABP has been associated with macrophage phenotype modulation under inflammatory conditions, which is significant in AMI pathophysiology^50^. In patients with AMI, IVNS1ABP expression decreases following mechanical reperfusion and stenting, with downregulation persisting up to one year post-AMI, highlighting its potential in early diagnosis and intervention^50,51^. This suggests that IVNS1ABP may play a role in the early phase of AMI, potentially serving as a biomarker for early prediction and treatment. However, the mechanisms underlying IVNS1ABP’s effects and downstream regulatory pathways in AMI warrant further investigation. Our study also identified IVNS1ABP as a risk factor for AMI.

PLAUR, also known as u-PAR, is a key regulator of cell surface fibrinogen activation, influencing numerous physiological and pathological processes, including colon cancer metastasis and progression^52,53^. PLAUR expression has been linked to the advancement of atherosclerotic lesions, with studies demonstrating that PLAUR expression in macrophages correlates with lesion progression in mouse models of atherosclerosis. Additionally, PLAUR deficiency or administration of soluble PLAUR can reduce atherosclerotic lesions in mice^54^, suggesting PLAUR as a potential therapeutic target for HLP-associated atherosclerosis. PLAUR is also involved in immune modulation, which may impact inflammatory processes in cardiovascular diseases^55^. Our findings indicate that PLAUR could be a risk factor for AMI. ROC analysis was subsequently conducted to evaluate the diagnostic value of the three HRGs and LRGs in AMI. The AUC values for PLAUR, QKI, and IVNS1ABP in the training set were 0.773, 0.933, and 0.807, respectively, indicating that each gene has strong diagnostic potential for AMI. Integrating these three genes into a nomogram model resulted in an AUC of 0.924, confirming the successful construction of the nomogram and the combined diagnostic utility of these biomarkers.

In this study, GSEA enrichment analysis of the three biomarkers (PLAUR, QKI, and IVNS1ABP) revealed significant associations with pathways including lipid and atherosclerosis, FoxO signaling, TNF signaling, apoptosis, complement and coagulation cascades, platelet activation, HIF-1 signaling, oxidative phosphorylation, and VEGF signaling. In AMI, the complex interplay between inflammation, energy metabolism, platelet function, and lipid metabolism is central to disease pathogenesis. Plaque rupture in coronary arteries exposes subendothelial tissue, initiating an inflammatory cascade. Cytokines such as interleukins (IL-1 and IL-6) and tumor necrosis factor-alpha (TNF-α) are released, further promoting inflammation and recruiting immune cells like macrophages and neutrophils to the injury site, thereby contributing to plaque progression and destabilization^56^. Myocardial ischemia, resulting from reduced blood flow and oxygen supply, disrupts energy metabolism, as myocardial cells struggle to generate sufficient adenosine triphosphate (ATP) through oxidative phosphorylation. The metabolic shift to anaerobic glycolysis yields less ATP and accumulates lactate, further impairing cardiac function. Under these hypoxic conditions, the hypoxia-inducible factor-1 (HIF-1) signaling pathway activates to restore oxygen homeostasis and support energy production^57^. Platelets, once activated, undergo shape change, release granules containing pro-inflammatory and pro-thrombotic factors, and aggregate to form a platelet plug. Interactions between platelets and damaged endothelium, combined with coagulation cascade activation, lead to rapid thrombus formation, intensifying the ischemic event^58^. Consequently, antiplatelet therapy is a cornerstone of AMI treatment, aimed at preventing additional thrombus formation. Dyslipidemia, characterized by elevated low-density lipoprotein cholesterol (LDL-C) and triglycerides, accelerates plaque formation. LDL-C particles can oxidize, forming oxidized LDL (ox-LDL), which is then engulfed by macrophages to form foam cells, advancing atherosclerotic plaques. This process ultimately contributes to AMI by triggering inflammation and thrombus formation^59^.

The interactions among inflammation, lipid metabolism, energy deprivation, and platelet activation are highly complex. Inflammation can drive further lipid accumulation and plaque growth, while energy deprivation may intensify platelet activation and coagulation. Additionally, the progression of atherosclerosis creates ischemic conditions that further disrupt energy metabolism in cardiac muscle^37^. Understanding these intricate interactions is vital for developing targeted AMI therapies. Anti-inflammatory drugs can help regulate immune responses, antithrombotic agents can prevent thrombus formation, and lipid-lowering therapies can address dyslipidemia, thereby stabilizing or even reversing atherosclerotic plaques. Strategies to improve energy metabolism, such as enhancing glucose uptake or promoting angiogenesis, are also being explored as potential therapeutic approaches. In summary, PLAUR, QKI, and IVNS1ABP may exert significant effects on AMI by modulating pathways related to inflammation, energy metabolism, platelet function, and lipid metabolism. This insight offers a fresh perspective on the potential of these biomarkers as therapeutic targets in AMI management.

Comparative analysis of PARG expression between AMI and control samples revealed that 12 of the 94 platelet activity-related genes were significantly differentially expressed. Specifically, SLC7A11, VAV1, DGKI, P2RX1, and CLIC1 showed increased expression in AMI samples, while CD40, ENTPD2, CD40LG, FIBP, TSPAN32, DGKE, and LCK were significantly downregulated. CLIC1 plays a critical role in platelet function—such as activation, thrombogenesis, and vasculogenesis—by interacting with integrins during cell adhesion, making it essential for maintaining platelet homeostasis and vascular integrity^60^. Our findings further underscore the significance of CLIC1 in AMI, with PLAUR showing the strongest positive correlation with CLIC1 and the strongest negative correlation with LCK, suggesting that PLAUR may contribute to AMI progression by enhancing platelet activation.

CD40LG, also known as CD40 ligand (CD40L), is integral to atherosclerosis development, impacting immune responses and arterial wall components. Platelet-expressed CD40LG can drive platelet activation and aggregation, fostering clot formation, a critical process in AMI^61,62^. Given that QKI emerged as a protective factor in this study and exhibited the strongest negative correlation with CD40LG, it is plausible that QKI may inhibit platelet activation and mitigate AMI risk by downregulating CD40LG expression. Overall, this analysis illuminates the potential roles of these PARGs in AMI, emphasizing the intricate interactions among genes involved in platelet activation, inflammation, and AMI pathophysiology. A deeper understanding of these relationships could inform the development of targeted therapies aimed at modulating platelet-related pathways in AMI.

Subsequent analysis identified 8 out of 84 TFs that commonly regulate all three biomarkers, underscoring their potential significance in MI progression. NANOG, for instance, is essential for cardiovascular phenotypes and survival, primarily through VEGF/VEGFR2 signaling activation^63^, while SMAD3 is central to TGF-β-induced cardiac fibrosis^64^. The lncRNA-miRNA-mRNA network comprised 21 differentially expressed circRNAs (DEcircRNAs), 11 differentially expressed miRNAs (DEmiRNAs), and 106 differentially expressed mRNAs (DEmRNAs). Among these, metastasis-associated lung adenocarcinoma transcript 1 (LncRNA MALAT1), a long non-coding RNA implicated in various biological processes^65–67^, functions as a molecular sponge for miR-106a-5p, sequestering it and thereby relieving its inhibitory effect on downstream targets^68^. One such target is QKI, a regulator of alternative splicing and RNA processing^68^. This interaction suggests that MALAT1 reduces miR-106a-5p availability, leading to increased QKI expression, which may, in turn, inhibit cardiomyocyte apoptosis. The regulatory interplay between MALAT1, miR-106a-5p, and QKI establishes a complex network that presents therapeutic potential for alleviating cardiac apoptosis in AMI. Elucidating these molecular interactions is essential for developing targeted strategies to address cardiovascular complications associated with AMI.

Our study identified piperlongumine as a potential therapeutic agent targeting the three biomarkers. Piperlongumine, a natural alkaloid derived from Piper longum L., exhibits a wide range of pharmacological activities, including antitumor, antiangiogenic, antiplatelet aggregation, anti-atherosclerotic, antidiabetic, and antibacterial effects^69^. Its antitumor properties are primarily attributed to its ability to elevate reactive oxygen species levels and modulate signaling pathways related to cell growth, survival, and apoptosis^70^. Furthermore, piperlongumine has shown promise in mitigating vascular remodeling in hypoxic pulmonary hypertension by regulating autophagy^71^. Studies have also demonstrated its effectiveness in suppressing rabbit platelet aggregation induced by various agents^72^ and preventing atherosclerotic plaque formation by reducing NF-κB activation^73^. These findings suggest that piperlongumine may hold therapeutic value not only as an anti-tumor agent but also in managing platelet activation and atherosclerosis, thereby underscoring its potential role in cardiovascular protection. Thus, piperlongumine could emerge as a targeted drug to inhibit AMI progression and potentially improve prognosis.

This research provides insights into the expression profiles and molecular pathways of LRGs and HRGs in AMI, laying the groundwork for precise diagnosis and the identification of novel therapeutic targets. However, clinical application of these bioinformatics findings requires validation through larger sample sizes. Additionally, further exploration and verification of these biomarkers’ roles in AMI through cell-based and in vivo studies will be essential to confirm their potential as therapeutic targets.

## Data Availability

All data produced in the present study are available upon reasonable request to the authors.

## Acknowledgments

We would like to express our sincere gratitude to all individuals and organizations who supported and assisted us throughout this research.

## Sources of Funding

This study was supported by Grants from the National Natural Science Foundation of China (82360077, 82460065), 535 Talent Project of First Affiliated Hospital of Kunming Medical University (2024535Q03), Yunnan Fundamental Research Projects (202201AU070063), Union Foundation of Yunnan Provincial Science and Technology Department and Kunming Medical University (202201AY070001-082).

## Disclosures

None.

## Ethical Approval and Informed Consent

I certify that this study was a part of certain research titled [The mechanism of let-7b mediated platelet autophagy in regulating platelet activation and thrombosis of ACS], which has been approved by the institutional review board (IRB) of the First Affiliated Hospital of Kunming Medical University. The approval number and date of approval are as follows: [2024L84] and [March 30, 2024].

Participants in this study were provided with a clear and understandable explanation of the research objectives, procedures, potential risks, and benefits. They were informed that their participation is voluntary and that they have the right to withdraw from the study at any time.

Participants were given the opportunity to ask questions and provided written informed consent prior to their involvement in the study.

## Non-standard abbreviations and acronyms

Abbreviations: Full name in English

HLP: Hyperlipidemia

TFs: Transcription factors

PARGs: Platelet activation-related genes

RF: Random forest

LASSO: Least absolute shrinkage and selection operator

LOO: Leave-One-Out

OR: Odds ratio

LD: Linkage disequilibrium

IVW: Inverse variance weighted

IEU: Integrative epidemiology unit

eQTL: Expression quantitative trait loci

LRGs: Lipophagy-related genes

DE-LRGs: Differentially expressed LRGs

GO: Gene ontology

FC: Fold change

PARGs: Platelet activation-related genes

ROC: Receiver perating characteristic

WGCNA: Weighted gene co-expression network analysis

CI: Confidence interval

OR: Odds ratio

RT-qPCR: Reverse transcription quantitative polymerase chain reaction

SNPs: Single nucleotide polymorphisms

ECG: Electrocardiogram

AUC: Area under the curve

HRGs: HLP-related genes

GEO: Gene expression omnibus

MI: Myocardial infarction

MSigDB: Molecular signatures database

RCTs: Randomized controlled trials

HRGs: HLP-related genes

MR: Mendelian randomization

AMI: Acute myocardial infarction

## Supplemental Material

Tables S1-S4

Figure S1-S4

## Notes

### Competing Interest Statement

The authors have declared no competing interest.

### Funding Statement

This study was funded by Grants from the National Natural Science Foundation of China (82360077, 82460065), 535 Talent Project of First Affiliated Hospital of Kunming Medical University (2024535Q03), Yunnan Fundamental Research Projects (202201AU070063), Union Foundation of Yunnan Provincial Science and Technology Department and Kunming Medical University (202201AY070001-082).

### Author Declarations

IRB of the First Affiliated Hospital of Kunming Medical University gave ethical approval for this work.

